# Rapid spread and high impact of the Variant of Concern P.1 in the largest city of Brazil

**DOI:** 10.1101/2021.04.10.21255111

**Authors:** Gabriela Rodrigues Barbosa, Luiz Vinicius Leão Moreira, Alberto Fernando Oliveira Justo, Ana Helena Perosa, Ana Paula Cunha Chaves, Mariana Sardinha Bueno, Luciano Kleber de Souza Luna, Danielle Dias Conte, Joseane Mayara Almeida Carvalho, Janesly Prates, Patricia Sousa Dantas, Klinger Soares Faico-Filho, Clarice Camargo, Paola Cristina Resende, Marilda Mendonça Siqueira, Nancy Bellei

**Author notes:** Corresponding author Gabriela Rodrigues Barbosa, Pedro de Toledo St, 781 - Vila Clementino, São Paulo, SP, Brazil, (ext 2222).

## Abstract

First in Manaus in the Brazilian Northern region, the Variant of Concern P.1 traveled 3800 kilometers southeast to endanger Sao Paulo contributing to the collapse of the health system. Here, we show evidence of how fast the VOC P.1 has spread in the most populated city in South America.

---

The COVID-19 has reached over 131 million cases around the world, with more than 2.8 million of death, according to the World Health Organization (1). As the SARS-CoV-2 cases continue to emerge globally, variants of concern (VOC) and variants of interest (VOI) have been described (2). The variants share important points of mutations at the receptor-binding domain (RBD) of the Spike protein, which might increase the transmissibility of COVID-19 and promote escape from neutralizing antibodies (3).

Brazil is currently the epicenter of COVID-19, with more than 13 million confirmed cases of April 2021(1). Two variants, VOC P.1 and VOI P.2, evolved from lineage B.1.1.28, have taken over the scene since late 2020 in the country (4,5). The VOC P.1 was first detected in January 2021 in Japanese travelers returning from Manaus and was responsible for the second wave in Amazonas in late November 2020 (5). In October 2020, the VOI P.2 was reported in Rio de Janeiro and was estimated to have emerged in late August 2020 (5,6). Both variants have been associated also with reinfection cases (6).

In a recent investigation (data not published) of the circulating variants in São Paulo city, during the first week of March, 64.4% of samples were identified as P.1. The investigation of lineages not only contributes to epidemiological surveillance but also provides a better comprehension of the spread and circulation of variants, allowing the association to clinical outcomes and response to vaccines (7). In this sense, we aimed to investigate the spread of P.1 and P.2 variants in samples from hospitalized patients (HP) and healthcare workers (HCW) attended in a university hospital in São Paulo city. This study was conducted in compliance with institutional guidelines, approved by the Ethics Committee of São Paulo Federal University (CEP/UNIFESP n. 29407720.4.0000.5505).

From March 1^st^ to March 15^th^, 427 nasopharyngeal samples were collected from 245 HP and 125 from HCW outpatients (25.5% and 23.2% of positivity rate, respectively). We then selected 60 samples with Ct value ≤30 (38 samples from HP, and 22 from HCW). All HCW presented only mild symptoms and did not need hospitalization.

Of the 60 selected samples, 52 whole genome sequences were generated (30 from HP and 22 from HCW) following the sequencing protocol using the Illumina MiSeq platform and the analysis pipeline described by Resende et al (8). The SARS-CoV-2 lineages were classified by the PANGO lineages nomenclature (9). Genome sequences generated have been deposited at the EpiCoV database on GISAID (https://www.gisaid.org/) under accession numbers EPI_ISL_1464630 to EPI_ISL_1464677.

Of the 52 sequenced samples, 44 (84.4%) were identified as VOC P.1; 5 (9.2%) as VOI P.2; 1 (1,9%) as B.1.1.7, and 2 (3,8%) B.1.1.28 (Figure). The most notable variants circulating in the second wave, including B.1.1.7 (detected first in the United Kingdom) and B.1.1.351 (detected first in South Africa), and P.1, are related to an increase of transmissibility (2,10). Interestingly, the P.1 variant was first identified in the State of Amazonas, about 3,800 kilometers apart from São Paulo (5). It is evident that the P.1 variant prevailed during the first two weeks of March, showing a regular distribution among HP and HCW with no difference in terms of age, sex, vaccination, and outcome (Table). From the first to the second weeks of March, we observed a higher frequency of P.1 (78.6% and 91.7%, respectively). In this survey, only one sample from a HP was identified as VOC B.1.1.7. The other two samples were identified as B.1.1.28, a widely spread lineage during the first wave in Brazil.

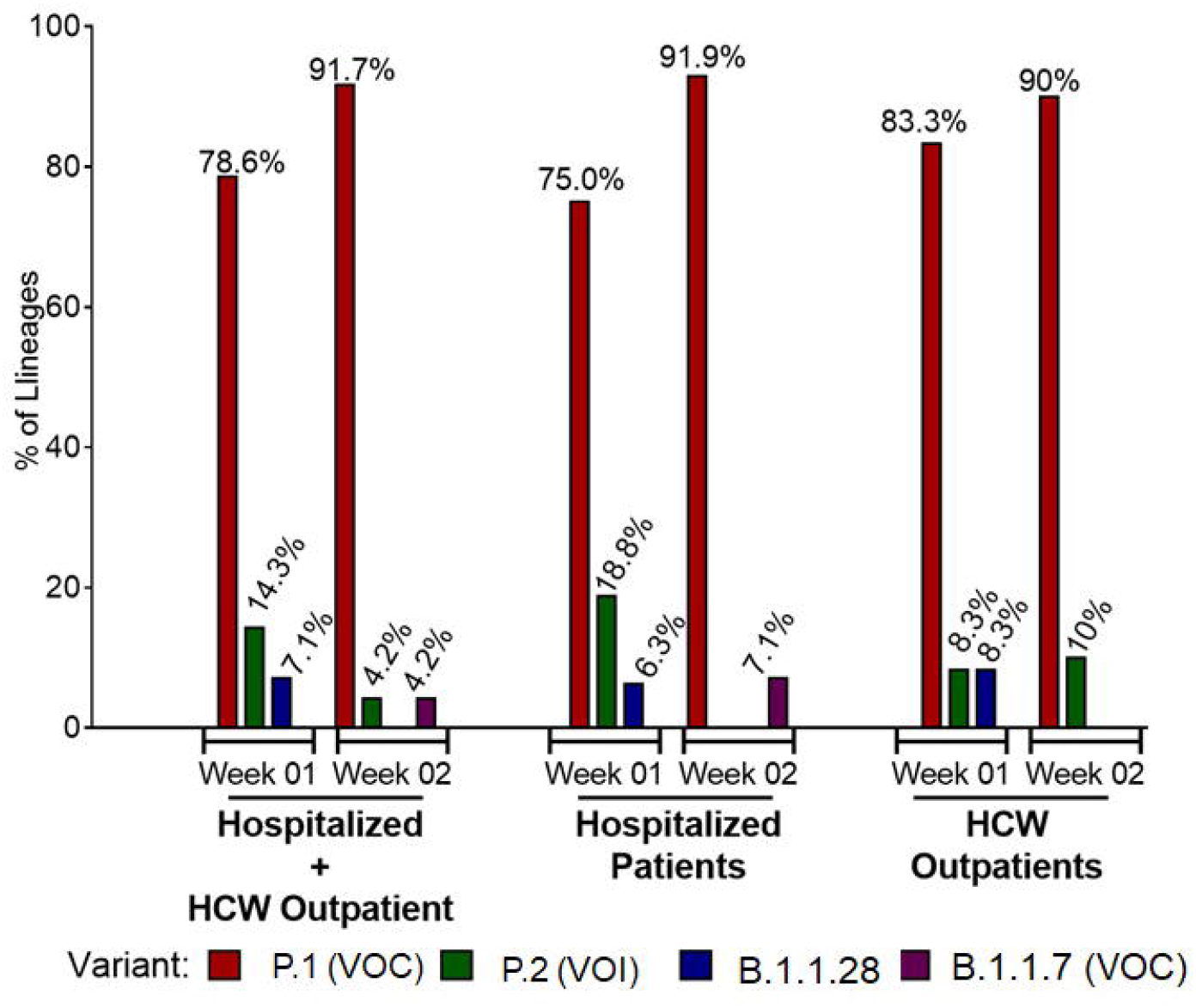

**Table.** Characteristics of individuals identified with the VOC P.1

|  | Hospitalized<br>Patients | HCW Outpatients |
| --- | --- | --- |
| **P.1** | **25** | **19** |
| Age (years) | 66 (33-86) | 29 (19-64) |
| Sex |  |  |
| Male | 14 | 5 |
| Female | 11 | 14 |
| Vaccinated | 1 | 10 |
| Death | 10 | - |
| Discharged | 15 | 19 |
| **Other variants** | **5** | **3** |
| Age (years) | 53 (36-71) | 23 (20-31) |
| Sex |  |  |
| Male | 2 | - |
| Female | 3 | 3 |
| Vaccinated | - | 3 |
| Death | 2 | - |
| Discharged | 3 | 3 |

There is a broad discussion about whether the available vaccines against SARS-CoV-2 will be less effective at preventing infection with the emerging variants (10). In this work, 14 samples (26.9%) of the 52 sequenced samples were from individuals that had received at least one dose of vaccine, ChAdOx1-S/nCoV-19 (n=2) or SINOVAC (n=26). Although they were vaccinated, they could not be considered immunized, regarding the days after vaccination.

Among the hospitalized patients, 19 (63%) were admitted to the intensive care unit, from which nine were discharged and ten died. Comparing the RT-PCR Ct values of all attended patients since the first wave, we did not observe any difference in the Ct mean values with those of P.1 (data not shown). May 2020 registered the peak of number of positive cases with a Ct mean of 23.6. Now, as of April 2021, we are facing a rise in the number of cases. However, the Ct mean was 24.9, which may indicate that the spread of P.1 does not contribute to an actual increase in the viral load.

There is still a need for more epidemiologic surveys to assure the role of the VOCs in transmission and escape to neutralizing antibodies. Our findings emphasize that the P.1 variant has spread widely throughout the country. Despite all actions of interventions such as the use of masks, physical distancing, flight travel reductions, and the currently established lockdown in São Paulo, the frequency rates of P.1 increased significantly in two weeks, evidencing its fast spread.

## Data Availability

The data that support the findings of this study are available in GISAID at https://www.gisaid.org/ reference number EPI_ISL_1464630 to EPI_ISL_1464677.

## Acknowledgements

The authors wish to thank all the health care workers and scientists who have worked hard to deal with this pandemic threat, in special the sequencing team from Laboratory of Respiratory Viruses and Measles, Oswaldo Cruz Institute and the Fiocruz COVID-19 Genomic Surveillance Network (http://www.genomahcov.fiocruz.br/).

This work was supported by grant 2020/11719-0, São Paulo Research Foundation (FAPESP) and Coordination for the Improvement of Higher Education Personnel CAPES.

